# Pathogenic autoantibody internalization in myositis

**DOI:** 10.1101/2024.01.15.24301339

**Authors:** Iago Pinal-Fernandez, Sandra Muñoz-Braceras, Maria Casal-Dominguez, Katherine Pak, Jiram Torres-Ruiz, Jon Musai, Stefania Dell’Orso, Faiza Naz, Shamima Islam, Gustavo Gutierrez-Cruz, Maria Dolores Cano, Ana Matas-Garcia, Joan Padrosa, Esther Tobías-Baraja, Gloria Garrabou, Iban Aldecoa, Gerard Espinosa, Carmen Pilar Simeon-Aznar, Alfredo Guillen-Del-Castillo, Albert Gil-Vila, Ernesto Trallero-Araguas, Lisa Christopher-Stine, Thomas E. Lloyd, Teerin Liewluck, Elie Naddaf, Werner Stenzel, Steven A. Greenberg, Josep Maria Grau, Albert Selva-O’Callaghan, Jose C. Milisenda, Andrew L. Mammen

## Abstract

**Objectives:** Myositis is a heterogeneous family of autoimmune muscle diseases. As myositis autoantibodies recognize intracellular proteins, their role in disease pathogenesis has been unclear. This study aimed to determine whether myositis autoantibodies reach their autoantigen targets within muscle cells and disrupt the normal function of these proteins.

**Methods:** Confocal immunofluorescence microscopy was used to localize antibodies and other proteins of interest in myositis muscle biopsies. Bulk RNA sequencing was used to study the transcriptomic profiles of 668 samples from patients with myositis, disease controls, and healthy controls. Antibodies from myositis patients were introduced into cultured myoblasts by electroporation and the transcriptomic profiles of the treated myoblasts were studied by bulk RNA sequencing.

**Results:** In patients with myositis autoantibodies, antibodies accumulated inside myofibers in the same subcellular compartment as the autoantigen. Each autoantibody was associated with effects consistent with dysfunction of its autoantigen, such as the derepression of genes normally repressed by Mi2/NuRD in patients with anti-Mi2 autoantibodies, the accumulation of RNAs degraded by the nuclear RNA exosome complex in patients with anti-PM/Scl autoantibodies targeting this complex, and the accumulation of lipids within myofibers of anti-HMGCR-positive patients. Internalization of patient immunoglobulin into cultured myoblasts recapitulated the transcriptomic phenotypes observed in human disease, including the derepression of Mi2/NuRD-regulated genes in anti-Mi2-positive dermatomyositis and the increased expression of genes normally degraded by the nuclear RNA exosome complex in anti-PM/Scl-positive myositis.

**Conclusions:** In myositis, autoantibodies are internalized into muscle fibers, disrupt the biological function of their autoantigen, and mediate the pathophysiology of the disease.

## INTRODUCTION

Myositis is a family of autoimmune disorders variably affecting multiple organs, including muscle, skin, lungs, and/or joints. Most myositis patients have autoantibodies targeting intracellular autoantigens. These autoantibodies define unique subsets of myositis patients and it has been hypothesized, but not proven, that they may be causally linked to the pathogenesis of the disease.^1-4^

Previous transcriptomic studies identified type I interferon as an important mediator of dermatomyositis pathogenesis.^5-8^ Moreover, specific transcriptomic markers were identified in autoantibody-defined myositis subgroups.^9^

Anti-Mi2-positive dermatomyositis patients have autoantibodies that recognize the nuclear MI2/NuRD complex, a transcriptional repressor. Recently, we found that in anti-Mi2-positive dermatomyositis, antibodies are deposited in the nuclei of the myofibers. We also demonstrated that muscle biopsies from anti-Mi2 positive patients express high levels of genes normally repressed by the Mi2/NuRD complex, suggesting that anti-Mi2 autoantibodies directly bind to their autoantigen and inhibit its function.^10^

In the current study, we analyzed muscle from myositis patients with a variety of myositis autoantibodies to determine whether antibodies accumulate in the same subcellular compartment as the relevant autoantigen. Moreover, we sought to identify transcriptomic profiles and other evidence of autoantigen dysfunction. Finally, we studied the transcriptomic profiles of cultured muscle cells following the internalization of antibodies obtained from myositis patients with different myositis autoantibodies

## METHODS

### Patients

Muscle tissue from patients who underwent diagnostic muscle biopsies and healthy volunteers at several centers specialized in neuromuscular diseases underwent bulk RNAseq analysis (Supplementary Table 1). A subset of these muscle biopsies was used for immunofluorescence studies. For antibody internalization experiments, immunoglobulin was purified from the serum of patients with myositis and healthy controls (Supplementary Methods).

### RNA sequencing

Bulk RNAseq was performed on frozen muscle biopsy specimens as previously described.^6,9-13^ Briefly, RNA was extracted with TRIzol. Libraries were either prepared with the NeoPrep system according to the TruSeq Stranded mRNA Library Prep protocol (Illumina, San Diego, CA) or with the NEBNext Poly(A) mRNA Magnetic Isolation Module and Ultra^™^ II Directional RNA Library Prep Kit for Illumina (New England BioLabs, ref. #E7490, and #E7760).

### Histopathology and immunofluorescence

Muscle biopsy sections processed for clinical purposes were stained for hematoxylin and eosin (H&E), oil-red O (ORO), Gömöri trichrome, CD56 (NCAM), membrane attack complex, NADH, and COX, and then microscopic images were digitized using a Leica Slide Scanner SCN400F.

For immunofluorescence studies, 10μm thick unfixed sections were incubated with primary and secondary antibodies (Supplementary Table 2). Images were obtained using a Leica SP8 high-resolution confocal microscope.

### Culture of differentiating human skeletal muscle myoblasts and treatment with different types of interferon

Normal human skeletal muscle myoblasts were cultured according to the protocol recommended by the supplier (Lonza). When 80% confluent, myoblasts were induced to differentiate into myotubes by replacing the growth medium with differentiation medium (DMEM-F12 [Lonza, ref. 12-719F] supplemented with 2% horse serum [Gibco, ref. 16050-122], insulin-transferrin-selenium [Gibco, ref. 41400-045], and penicillin-streptomycin-L-glutamine [Gibco, ref. 10378-016]). Cells were harvested before differentiation and then daily after differentiation for 6 days.

To examine the effect of different types of interferon on gene expression we treated the cells daily with 100 U/uL and 1000 U/uL of IFNA2a (R&D, ref. 11100-1), and IFNB1 (PeproTech, ref. 300-02BC), respectively, for 7 days. Then, the cells were harvested for RNA extraction and RNA sequencing.

### Electroporation of antibodies into human muscle cells

Human immunoglobulins were purified and concentrated from serum using protein G Agarose (Millipore, ref. 16-266) and the Amicon Pro Purification System (Millipore, ref. ACS500024) with a 30kDa molecular weight cutoff Amicon Ultra Centrifugal Filter (Millipore, ref. UFC503024).

Normal human skeletal muscle myoblasts were cultured and nucleofected with purified immunoglobulins according to the protocol recommended by the supplier (Lonza) and using the P5 Primary Cell 4D-Nucleofector™ X Kit L (Lonza, ref. V4XP-5024). Nucleofected cells were plated in differentiation medium for different numbers of days and harvested for RNA extraction and subsequent RNA sequencing 24 hours after unless otherwise indicated.

### Statistical and bioinformatic analysis

For RNAseq analysis, sequencing reads were demultiplexed using bcl2fastq/2.20.0 and preprocessed using fastp/0.21.0. The abundance of each gene was determined using Salmon/1.5.2. Counts were normalized using the Trimmed Means of M values (TMM) from edgeR/3.34.1 for graphical analysis. Differential expression was performed using limma/3.48.3. The Benjamini-Hochberg correction was used to adjust for multiple comparisons if appropriate. Pathway enrichment analysis employed a one-sided Fisher’s exact test.

To define the specific set of genes associated with each group of interest we calculated the intersection of the differentially overexpressed genes (q-value < 0.01) between the group of interest and each of the other comparator groups. Venn diagrams were used to represent graphically these analyses.

For immunofluorescence image analysis, individual muscle fibers were segmented using Cellpose 2.1.1^14^ using the neural network model “Cytoplasm 2.0” and the intensity of human immunoglobulin and MX1 in individual fibers was quantified using ImageJ2 2.9.0. Fibers smaller than 25μm of Feret’s diameter were excluded from the analysis.

## RESULTS

### Antibodies are deposited within myofibers in patients with myositis autoantibodies

Immunoglobulin G staining revealed that antibodies were deposited inside the muscle fibers of patients with each of the different myositis-specific autoantibodies. Myofibers of patients with anti-HMGCR, anti-SRP, anti-MDA5, and anti-Jo1 autoantibodies, all of which recognize cytoplasmic autoantigens, displayed a cytoplasmic pattern of immunoglobulin deposition. In contrast, biopsies from those with anti-Mi2 autoantibodies, which target nuclear proteins, and those with anti-PM/Scl autoantibodies, targeting proteins in the nucleolus, exhibited predominantly nuclear and nucleolar immunoglobulin deposition, respectively. Interestingly, although anti-NXP2 and anti-TIF1g autoantibodies targeting nuclear antigens, muscle fibers from these patients had a predominant cytoplasmic pattern of immunoglobulin deposition (Figure 1, Supplementary Figures 1-20). Of note, in patients with anti-NXP2 autoantibodies, its autoantigen was aberrantly localized in the cytoplasm, as will be discussed below.

**Figure 1.**
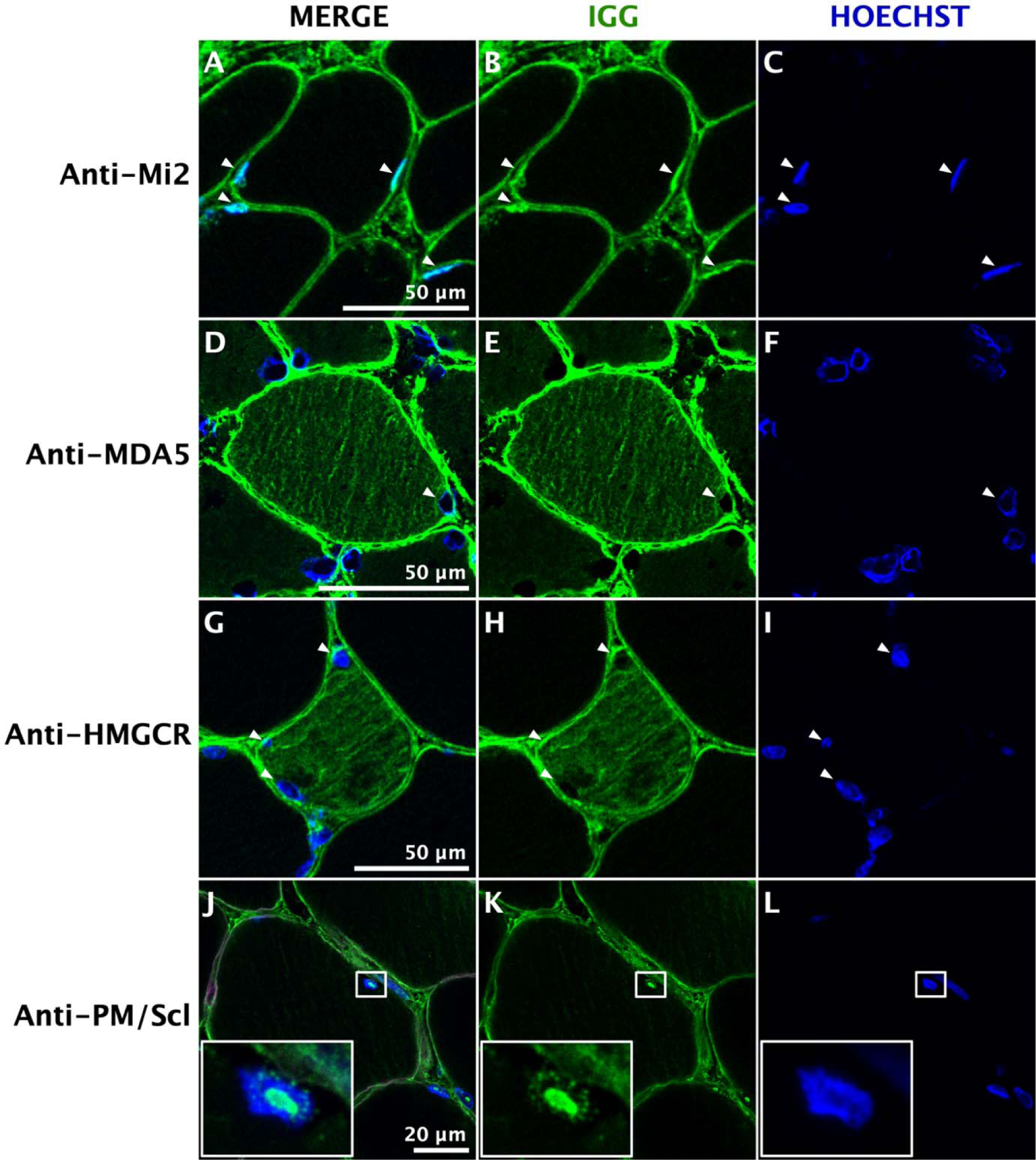
Immunoglobulin localization in myositis muscle. Confocal immunofluorescence of human immunoglobulin (IgG) in different autoantibody-defined types of myositis shows antibody deposition in the nuclei (white arrowheads) of muscle fibers from anti-Mi2-positive patients (A-C); the cytoplasm of anti-MDA5-positive (D-F) and anti-HMGCR-positive (G-I) patients; and the nucleoli of anti-PM/Scl-positive patients (J-L). The square box contains a higher magnification image of one nucleus, showcasing the nucleolar pattern of IgG.

### IFN1 overexpression in dermatomyositis muscle is driven by IFNB1 and correlates with antibody internalization

Type I interferon-stimulated genes are highly overexpressed in dermatomyositis muscle tissue (Figures 2 and 3).^5,6,15^ The function of certain myositis autoantigens is to regulate the expression of single type-I interferon proteins (e.g. NXP2^16^ and TIF1g^16,17^ inhibit IFNB1 expression). To further clarify the relationship between myositis autoantibodies and the interferon pathway, we first had to define the specific type I interferon responsible for upregulating type I interferon-stimulated genes in myositis muscle.

**Figure 2.**
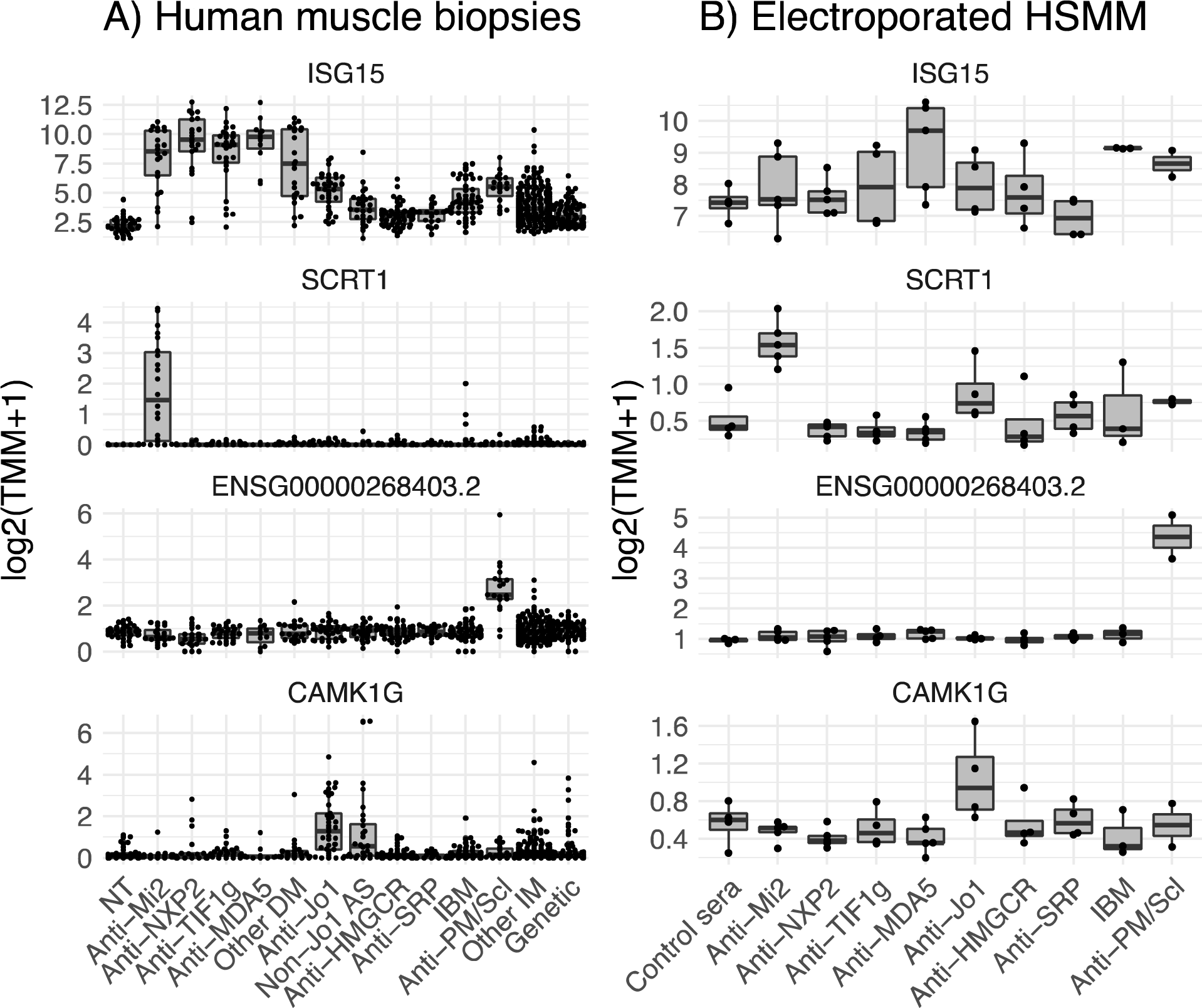
Representative genes expressed in muscle biopsies and cultured myoblasts electroporated with antibodies from myositis patients. Muscle biopsies from all dermatomyositis subgroups (A) and cultured muscle cells with internalized antibodies from 3/5 anti-MDA5-positive patients (B) overexpressed ISG15, a representative IFNB1-stimulated gene. Anti-Mi2-positive muscle biopsies (A) and cultured cells electroporated with antibodies from anti-Mi2-positive patients (B) overexpressed SCRT1, a representative gene from the anti-Mi2-specific gene set. Anti-PM/Scl-positive muscle biopsies (A) and cultured cells electroporated with antibodies from anti-PM/Scl-positive patients (B) overexpressed ENSG00000268403.2, a representative long non-coding RNA from the anti-PM/Scl-specific gene set. Muscle biopsies from anti-synthetase patients (A) and cultured cells electroporated with antibodies from anti-Jo1 patients (B) overexpressed CAMK1G, a member of the anti-Jo1-specific gene set. NT: histologically normal muscle biopsies; DM: dermatomyositis; AS: antisynthetase syndrome; IBM: inclusion body myositis; IM: inflammatory myopathies.

**Figure 3.**
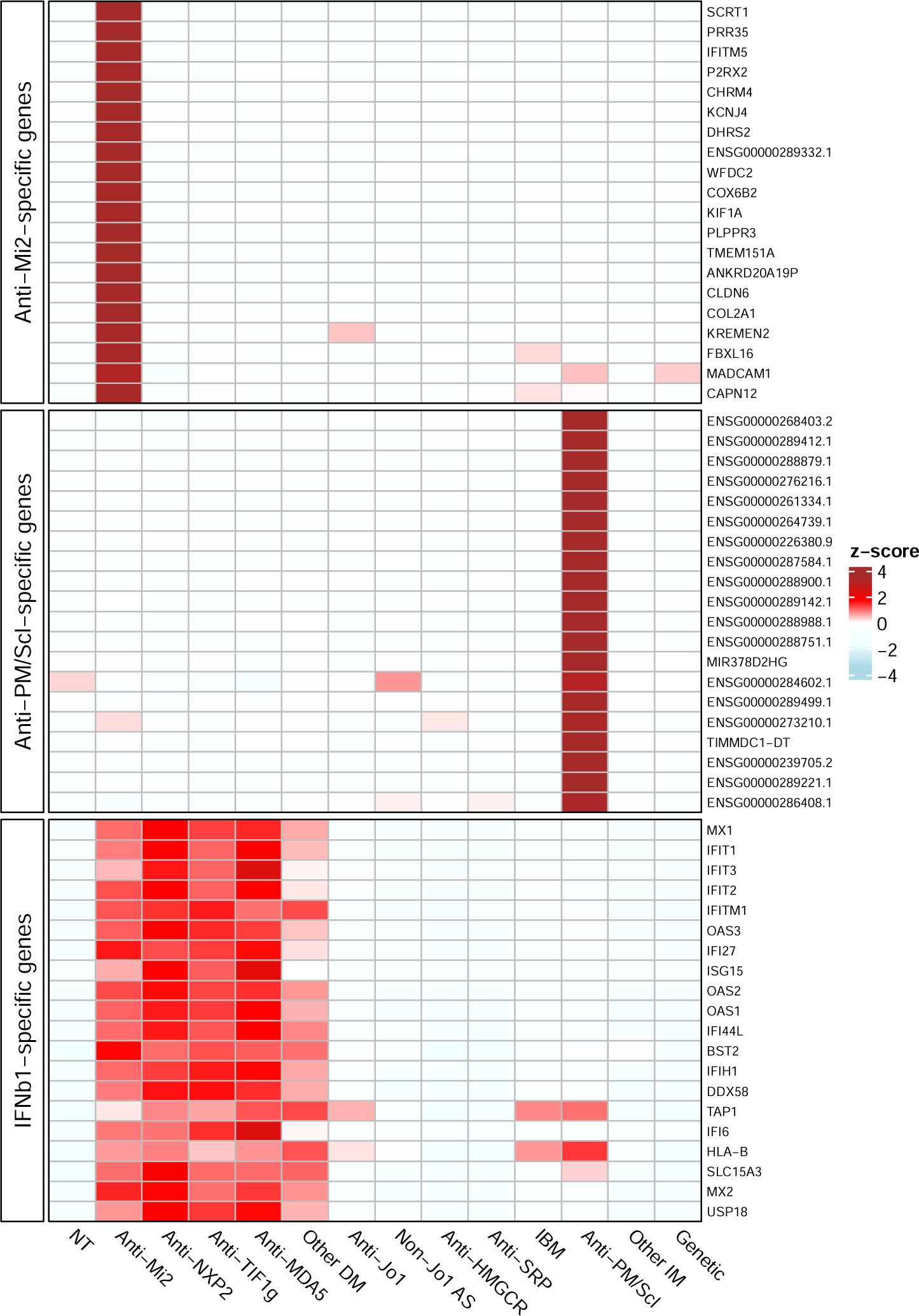
Gene expression patterns in muscle biopsies. Standardized expression levels (Z-scores of the median) are shown for the top 20 anti-Mi2-specific genes, the top 20 anti-PM/Scl-specific genes, and the top 20 IFNB1-specific genes in muscle biopsies from patients with different types of myopathies and histologically normal muscle biopsies (NT). The set of IFNB1-specific genes was derived from RNAseq data of cultured human muscle cells treated with different type I interferons. DM: dermatomyositis; AS: antisynthetase syndrome; IBM: inclusion body myositis; IM: inflammatory myopathies.

Using our transcriptomic dataset, we found that IFNB1 is the predominant type I interferon detected in dermatomyositis muscle biopsies. Also, IFNB1 expression correlated with the expression of IFN1-inducible genes (e.g. MX1) in muscle biopsies from dermatomyositis patients (Supplementary Figures 21-22). Furthermore, in myofibers, we observed a positive correlation between the intensity of internalized immunoglobulin and MX1 immunostaining (Supplementary Figures 23-28). Finally, IFNB1 treatment stimulated its own expression in cultured differentiated human myoblasts (Supplementary Figure 29). This suggests the possibility that the overexpression of IFNB1 can persist through a self-sustaining loop and confirms the results from publicly available datasets suggesting the same phenomenon in other cell types (monocytes [GSE34627]^18^, and PBMCs [GSE16214]^19^).

### Internalization of antibodies from anti-Mi2 patients causes the derepression of Mi2/NURD-regulated genes

Anti-Mi2 autoantibodies bind to CHD components of the Mi2/NuRD complex,^20^ which is a transcriptional repressor.^21^

Here, using our expanded muscle biopsy RNAseq dataset, we defined anti-Mi2-specific genes by calculating the intersection of the differentially expressed genes (q-value cutoff < 0.01) between anti-Mi2-positive patients and each of the other comparator groups. This analysis revealed more than 100 genes (e.g. SCRT1) that are exclusively overexpressed in anti-Mi2 muscle biopsies (Figures 2 and 3, Supplementary Figures 30-32, Supplementary Table 3).^10^ Of note, we previously established that this anti-Mi2-specific gene set is highly enriched for genes known to be transcriptionally repressed by the Mi2/NuRD complex.

Internalization of purified antibodies from anti-Mi2-positive patients into cultured muscle cells induced the overexpression of the same set of anti-Mi2-specific genes that are observed in the muscle biopsies of dermatomyositis patients with anti-Mi2 autoantibodies (Figure 4, Supplementary Figures 33-35). Of note, incubation of purified antibodies without electroporation had no transcriptomic effect in any of the autoantibody groups that we studied (Supplementary Figure 36).

**Figure 4.**
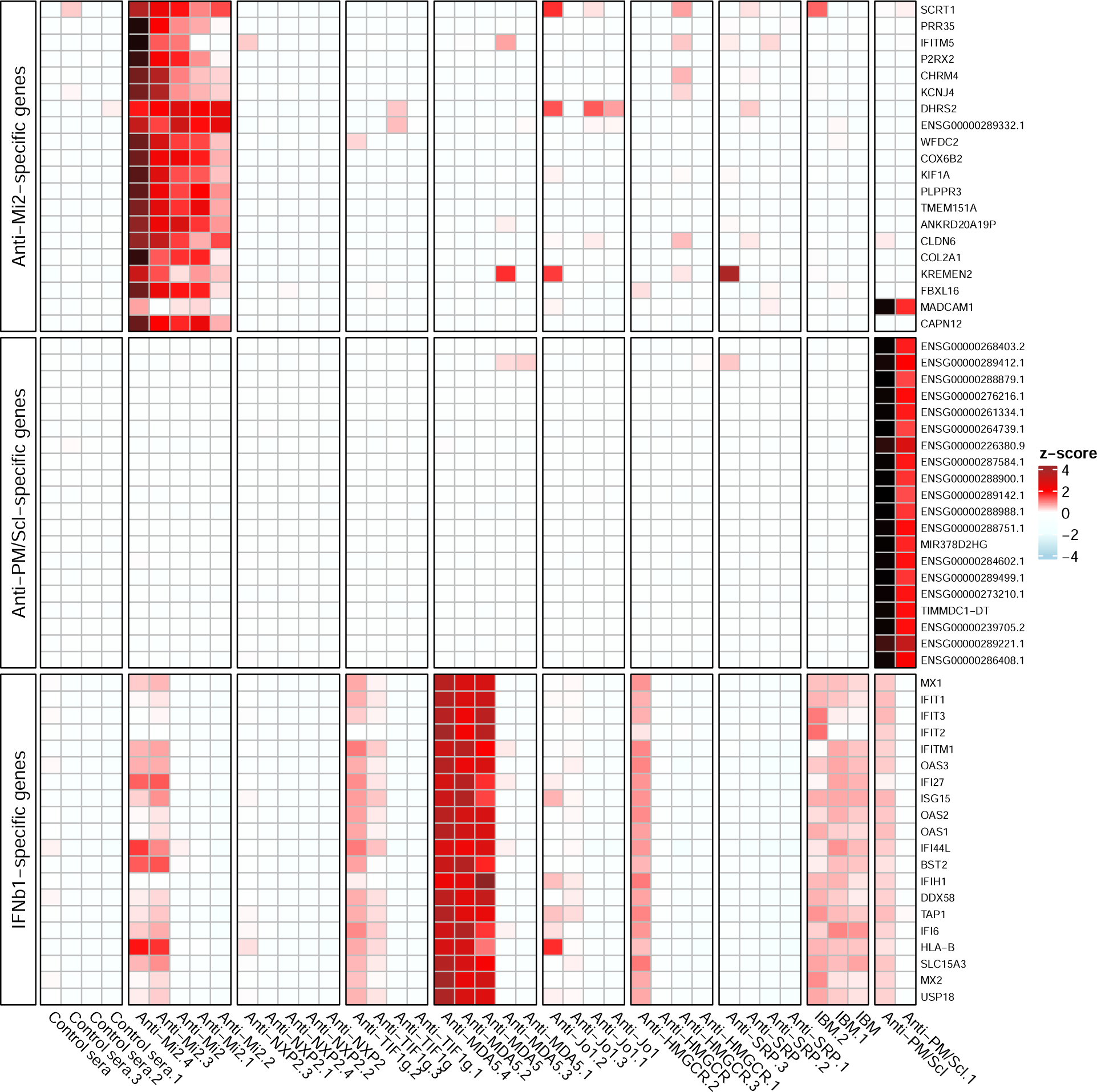
Gene expression patterns in cultured human myoblasts with internalized patient antibodies. Standardized expression levels (Z-scores) of the top 20 anti-Mi2-specific genes, the top 20 anti-PM/Scl-specific genes, and the top 20 IFNB1-specific genes are shown for cultured human myoblasts electroporated with purified antibodies from individual myositis patients and healthy controls. Anti-Mi2-specific genes are overexpressed predominantly in myoblasts electroporated with antibodies from anti-Mi2-positive patients. Anti-PM/Scl-specific genes are overexpressed exclusively in myoblasts electroporated with antibodies from anti-PM/Scl-positive patients. IFNB1-specific genes are most highly expressed in myoblasts electroporated with antibodies from 3 of 5 anti-MDA5-positive patients.

### Antibodies from anti-PM/Scl-positive patients disrupt the function of the nuclear RNA exosome complex

Anti-PM/Scl autoantibodies recognize EXOSC9 and EXOSC10, which are key components of the nuclear RNA exosome complex. The biological function of the exosome complex is to degrade various types of RNA, including long noncoding RNAs and divergent transcripts.^22,23^

We performed bulk RNAseq on 19 muscle biopsies from patients with anti-PM/Scl autoantibodies and compared them to the rest of the samples included in the study. We defined the set of anti-PM/Scl-specific genes, by calculating the intersection of the differentially expressed genes (q-value cutoff < 0.01) between anti-PM/Scl-positive patients and each of the other comparator groups (Supplementary Figures 37-39, Supplementary Table 4). This analysis identified 236 overexpressed RNAs and one underexpressed gene. Most overexpressed genes were long noncoding RNAs and divergent transcripts, suggesting a dysfunction of the nuclear RNA exosome complex exclusively in anti-PM/Scl-positive patients. Of note, only a single overexpressed gene was identified amongst all the other non-anti-Mi2 autoantibody groups (Supplementary Figures 40-41).

To verify that antibodies from anti-PM/Scl patients induce the expression of the anti-PM/Scl-specific gene set, we electroporated purified antibodies from anti-PM/Scl patients into human myoblasts. Indeed, this treatment induced the overexpression of the same set of anti-PM/Scl-specific genes that are observed in the muscle biopsies of anti-PM/Scl-positive patients (Figures 2, 4 and Supplementary Figures 42, 43).

### Antibodies from anti-MDA5 patients induce overexpression of IFNB1

Anti-MDA5 autoantibodies bind to the helicase domains of the MDA5 protein.^24^ MDA5 is primarily a cytoplasmic sensor of viral double-stranded RNA. Binding to double-stranded RNA activates the MDA5 protein, which ultimately induces the transcription of type I interferon.^25^

Internalization of purified immunoglobulin from 3 of 5 anti-MDA5 patients into human myoblasts induced a robust overexpression of IFNB1 and IFNB1-inducible genes (Figure 4, Supplementary Figures 35 and 44). This suggests the possibility that anti-MDA5 autoantibodies bind and activate MDA5.

### Internalization of antibodies from antisynthetase patients induces a transcriptional phenotype consistent with dysfunction of aminoacyl-tRNA synthetase

Anti-synthetase autoantibodies recognize members of the aminoacyl-tRNA synthetase family of proteins. These enzymes load the appropriate amino acid into its corresponding tRNA for eventual integration onto an elongating polypeptide. The most common of these autoantibodies recognize the histidyl-tRNA synthetase (anti-Jo1). It has been demonstrated that anti-Jo1 autoantibodies inhibit the function of its target protein *in vitro*.^26^

Previously, we reported that a set of genes including CAMK1G, EGR4, and CXCL8 are overexpressed in the muscle biopsies of patients with anti-Jo1 autoantibodies. In this study, we have expanded our cohort by including muscle biopsies from patients with non-Jo1 antisynthetase autoantibodies (e.g. anti-PL7 and -PL12). These tissue samples also overexpress the set of genes that were elevated in muscle biopsies from anti-Jo1-positive patients (Figure 2, Supplementary Figures 45-46, Supplementary Table 5).

However, this transcriptomic signature is weaker than those observed in muscle biopsies from anti-Mi2 or anti-PM/Scl-positive patients.

Internalization of purified immunoglobulin from anti-Jo1 patients into cultured human myoblasts induced overexpression of some of the same genes that we identified in patients with antisynthetase autoantibodies (Figure 2).

To test if this transcriptional program may be the consequence of aminoacyl tRNA-transferase dysfunction, we compared our gene set with a publicly available gene expression dataset from HepG2 human hepatoma cells treated with histidinol, an inhibitor of histidyl-tRNA synthetase.^27^ This dataset showed overexpression of genes specifically overexpressed in patients with the antisynthetase syndrome (e.g. EGR4, CXCL8). In addition, pathway enrichment analysis demonstrated a significant association between the genes in this dataset and those differentially expressed in patients with antisynthetase autoantibodies (p<0.001).

### Lipids accumulate in the muscle fibers of anti-HMGCR-positive patients

Anti-HMGCR autoantibodies recognize the rate-limiting enzyme of the cholesterol biosynthetic pathway. Recent studies demonstrated that mutations disrupting the enzymatic activity of HMGCR cause a genetic myopathy characterized by myofiber necrosis. This suggests the possibility that disruption of HMGCR by autoantibodies could lead to the same pathogenic abnormalities.^28,29^

Upon review of all available muscle biopsies, we noted accumulations of lipids in myofibers of patients with anti-HMGCR autoantibodies (Supplementary Figures 47-49). Specifically, 90% (18/20) of muscle biopsies from anti-HMGCR-positive patients had lipid accumulation compared to 8.3% (10/110) of patients with other types of myositis (p<0.001). As myofibers from patients under pharmacologic inhibition of HMGCR by statins may also have prominent lipid accumulations^30^, we hypothesize that autoantibody-mediated HMGCR dysfunction may induce the same effect by causing the accumulation of acetyl-CoA and the subsequent production of excess lipids.

Of note, anti-HMGCR myositis lacks a robust transcriptional signature and the only specific differentially expressed gene that we could identify was APOA4.^9^ However, internalization of antibodies from anti-HMGCR-positive patients into cultures myoblasts did not lead to an increase in APOA4 expression.

### In patients with anti-NXP2 autoantibodies, the target protein is aberrantly localized to the cytoplasm

Both NXP2^16^ and TIF1g^17^ proteins inhibit IFNB1 expression by binding to regulatory regions of this gene in the nuclei of cells.^16,17^

We could not find a reliable commercial anti-TIF1g antibody histochemical reagent. However, muscle biopsies from anti-NXP2 patients co-stained for NXP2 protein and immunoglobulin showed that both proteins were predominantly co-localized in the cytoplasm (Supplementary Figures 18-19), while NXP2 predominantly showed nuclear staining in non-NXP2 biopsies (Supplementary Figure 20).

The fact that the NXP2 protein is mislocalized to the cytoplasm suggests that the anti-NXP2 autoantibodies may sequester their autoantigen in the cytoplasm of muscle cells. This could cause increased expression of IFNB1 in the absence of inhibition by nuclear NXP2.

Electroporation of immunoglobulin from anti-NXP2 and anti-TIF1g patients into myoblasts did not activate the IFNB1 pathway. However, it should be noted that electroporation generates pores in the nuclear membrane, allowing antibodies to enter the cell nuclei. This would prevent the hypothesized cytoplasmic sequestration of NXP2 and TIF1g by their cognate autoantibodies.

## DISCUSSION

This study reveals two novel features of myositis pathogenesis. First, our immunofluorescence analyses demonstrate that myositis autoantibodies are internalized within living human muscle cells from patients with a variety of myositis autoantibodies. Second, using antibody internalization experiments along with transcriptomic analyses, we have shown that these internalized autoantibodies disrupt the normal function of their protein targets. Considering the essential roles of myositis autoantigens in cellular function and/or immunity (e.g. interferon production), our findings suggest that the autoantibody-mediated dysfunction of myositis autoantigens is likely to contribute to muscle damage and trigger the autoimmune response in patients with myositis (Figure 5).

**Figure 5.**
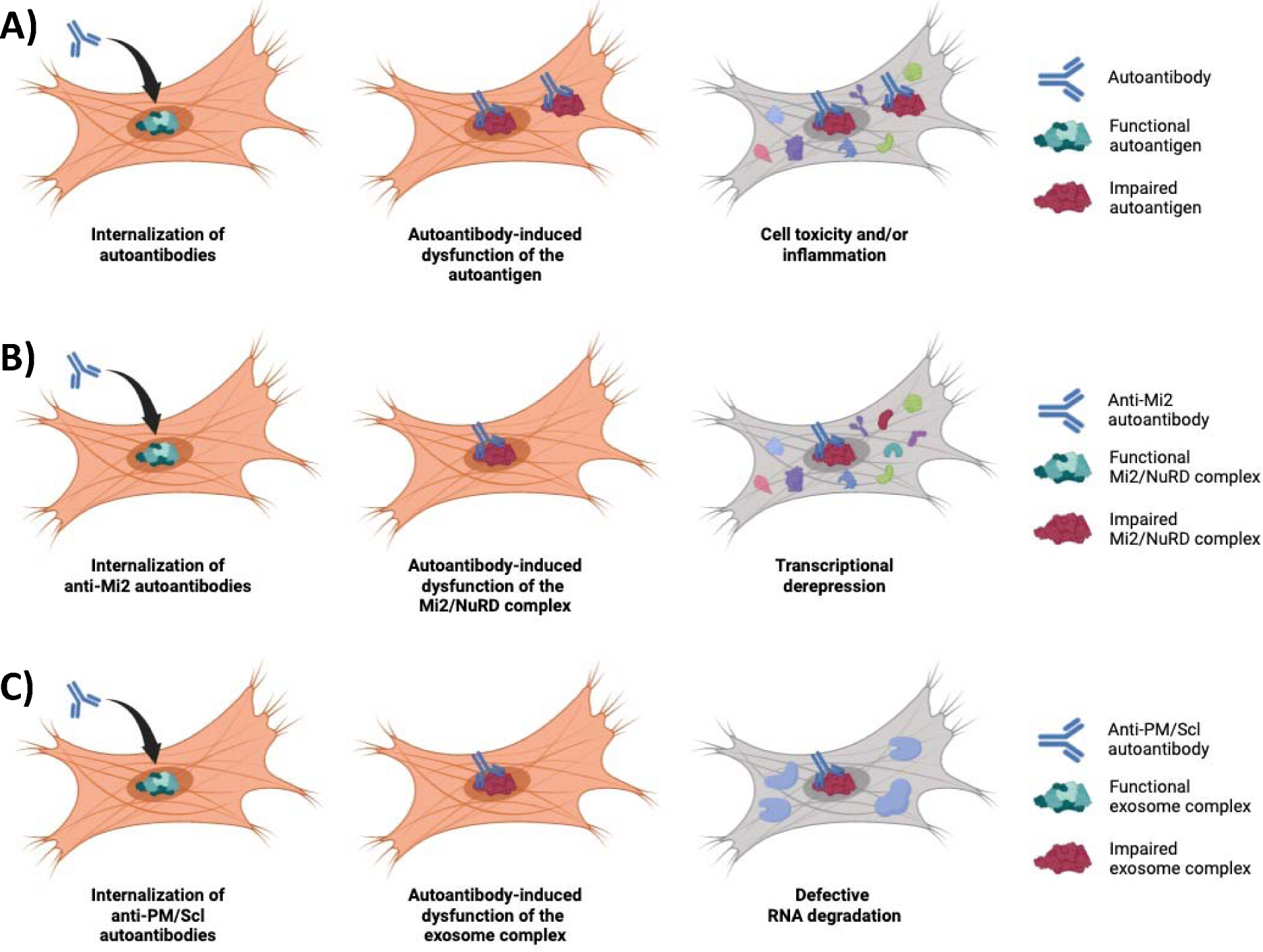
In myositis, autoantibodies are internalized into the muscle fibers, disrupting the normal biological function of their autoantigen, which mediates the pathogenesis of the disease (A). For instance, anti-Mi2 autoantibodies (B) interfere with the Mi2/NuRD complex inducing the derepression of more than 100 genes. Similarly, anti-PM/Scl autoantibodies (C) cause a dysfunction of the nuclear RNA exosome complex, impairing the normal degradation of various types of RNA.

In patients with autoantibodies targeting autoantigens involved in transcriptional regulation, notably those linked to the Mi2/NuRD complex in anti-Mi2 individuals or the exosome complex in patients with anti-PM/Scl autoantibodies, we observed transcriptomic effects consistent with dysfunction of their targets. Conversely, and as expected, muscle biopsies from patients with autoantibodies targeting proteins with no broad transcriptional functions, such as suppression of IFNB1 (TIF1g and anti-NXP2), interferon activation (MDA5), protein synthesis (aminoacyl-tRNA synthetases), protein trafficking (SRP), or lipid metabolism (HMGCR), exhibited less distinct transcriptional patterns. Nonetheless, we demonstrated evidence of amino acid deprivation in antisynthetase syndrome muscle biopsies, lipid dysregulation in muscle biopsies from anti-HMGCR-positive patients, and heightened interferon pathway activation in muscle biopsies from patients with anti-TIF1, anti-NXP2, and anti-MDA5 autoantibodies. These findings are also consistent with a disease mechanism in which myositis autoantibodies are pathogenic by modifying the normal function of their autoantigens (Figure 5).

Our investigation has several limitations. First, we only studied antibody internalization in muscle cells. Additional studies will be required to determine whether autoantibodies enter other cells within the muscle tissue, such as endothelial cells and fibroblasts, where they could also cause functional effects. Second, we did not study other organ systems frequently affected in myositis patients, such as the lungs or skin, to determine whether autoantibody internalization may play a pathophysiologic role in these locations. Furthermore, our study was limited to studying autoantibodies found in patients with myositis. As other systemic autoimmune diseases, including systemic sclerosis, vasculitis, and lupus are also associated with autoantibodies, the potential role of autoantibody internalization in the pathogenesis of these diseases remains a possibility. Further research exploring the potential pathologic role of autoantibody internalization in different cell types and tissues, as well as in different rheumatic diseases, will be required. Finally, although we show that myositis autoantibodies can enter living muscle cells and disrupt the normal function of their targets in either the cytoplasm or the nucleus, the mechanisms mediating autoantibody internalization remain to be elucidated.

In conclusion, our study shows that autoantibodies are deposited inside live muscle cells where they disrupt the normal function of their target proteins, contributing to the pathophysiology of this family of diseases. Beyond myositis muscle, this pathophysiologic mechanism may be relevant in other tissues and in other autoimmune diseases.

## Contributorship

All authors contributed to the development of the manuscript, including interpretation of results, substantive review of drafts, and approval of the final draft for submission.

## Data Availability

Any anonymized data not published within the article will be shared by request from any qualified investigator.

## Acknowledgments

None.

## Ethical approval information

All biopsies were from subjects enrolled in institutional review board (IRB)-approved longitudinal cohorts in the National Institutes of Health, the Johns Hopkins, the Clinic Hospital, the Vall d’Hebron Hospital, the Mayo Clinic, and the Charité-Universitätsmedizin Berlin.

